# Comparison of the clinical characteristics and outcomes of hospitalized adult COVID-19 and influenza patients – a prospective observational study

**DOI:** 10.1101/2020.06.29.20140632

**Authors:** Raija Auvinen, Hanna Nohynek, Ritva Syrjänen, Jukka Ollgren, Tuija Kerttula, Jarkko Mäntylä, Niina Ikonen, Raisa Loginov, Anu Haveri, Satu Kurkela, Kirsi Skogberg

## Abstract

**Objective:** We compared the clinical characteristics, findings and outcomes of hospitalized patients with coronavirus disease 2019 (COVID-19) or influenza to detect relevant differences.

**Methods:** From December 2019 to April 2020, we recruited all eligible hospitalized adults with respiratory infection to a prospective observational study at the HUS Jorvi Hospital, Finland. Influenza and SARS-CoV-2 infections were confirmed by RT-PCR. Follow-up lasted for at least 30 days from admission.

**Results:** We included 61 patients, of whom 28 were COVID-19 and 33 influenza patients with median ages of 53 and 56 years. Majority of both COVID-19 and influenza patients were men (61% vs 67%) and had at least one comorbidity (68% vs 85%). Pulmonary diseases and current smoking were less common among COVID-19 than influenza patients (5 [18%] vs 15 [45%], P=0.03 and 1 [4%] vs 10 [30%], P=0.008). In chest x-ray at admission, ground-glass opacities and consolidations were more frequent among COVID-19 than influenza patients (19 [68%] and 7 [21%], P < 0.001). Severe disease and intensive care unit (ICU) admission occurred more often among COVID-19 than influenza patients (26 [93%] vs 19 [58%], P=0.003 and 8 [29%] vs 2 [6%], P=0.034). COVID-19 patients were hospitalized longer than influenza patients (6 days [IQR 4-21] vs 3 [2-4], P<0.001).

**Conclusion:** Bilateral ground-glass opacities and consolidations in chest X-ray may help to differentiate COVID-19 from influenza. Hospitalized COVID-19 patients had more severe disease, required longer hospitalization and were admitted to ICU more often than influenza patients, which has important implications for public health policies.

## Introduction

Coronavirus disease 2019 (COVID-19) pandemic caused by severe acute respiratory syndrome coronavirus 2 (SARS-CoV-2) is currently causing such dramatic effects on health, economy and society as a whole that seasonal influenza, disruptive as it is, may seem to pale in comparison. However, several influenza pandemics have swept the globe and pandemic preparedness plans now used for COVID-19 are based on lessons from influenza. Understanding the differences in their epidemiology and clinical course is crucial for planning appropriate control measures for future COVID-19 mitigation.

COVID-19 first emerged in Wuhan, China, in December 2019 and spread to a worldwide pandemic by March 2020 [1-4]. By 24 June 2020, 9 129 146 cases and 473 797 deaths had been reported globally [5]. In comparison, the pandemic A(H1N1) virus caused an estimated 151 700 – 575 500 respiratory and cardiovascular deaths during the first 12 months of the latest influenza pandemic of 2009-2010 while seasonal influenza is estimated to be responsible for 291 243–645 832 respiratory deaths annually [6,7].

The clinical presentations of COVID-19 and influenza vary from mild respiratory tract infection to severe viral pneumonia leading to acute respiratory distress syndrome (ARDS) and death [3,8,-12]. In the largest study on COVID-19 to date, 80.9% of cases were considered mild, 13.8% severe and 4.7% critical [8]. A proportion of SARS-Cov-2 infections appear to be asymptomatic yet contagious [13-15]. Transmission can occur 1-2 days before symptom onset in both influenza and COVID-19 [15,16]. Fever and cough are the most common symptoms in both infections while fatigue, myalgia, headache and dyspnea may be present [3,8,-10,12,17,18]. Gastrointestinal (GI) symptoms occur in almost 20% of COVID-19 and 3-31% of influenza patients [19, 20]. Olfactory and taste disorders are associated with COVID-19 [21].

Well-known high-risk groups of influenza include elderly, pregnant women, under 2-year-old children and people with pre-existing comorbidities [12,18]. Established risk factors for the development of severe infection, ARDS and death in COVID-19 include older age, male sex and comorbidities [8,10,11,17,22,23]. While 80% of deaths occurred among under 65-year-olds during the 2009 A(H1N1) influenza pandemic, the mortality caused by COVID-19 has been highest among 80+-year-olds [6,8].

In Finland, the first cases of COVID-19 appeared in the HUS area in late February (Figure 1). Our prospective population-based influenza study was already recruiting hospitalized adult patients with severe acute respiratory infection (SARI), which gave us the opportunity to enroll COVID-19 patients since the beginning of the outbreak. We compared the clinical characteristics and outcomes of hospitalized adult COVID-19 and influenza patients to provide information for clinical diagnostics and public health strategies used in COVID-19 mitigation.

**Figure 1.**
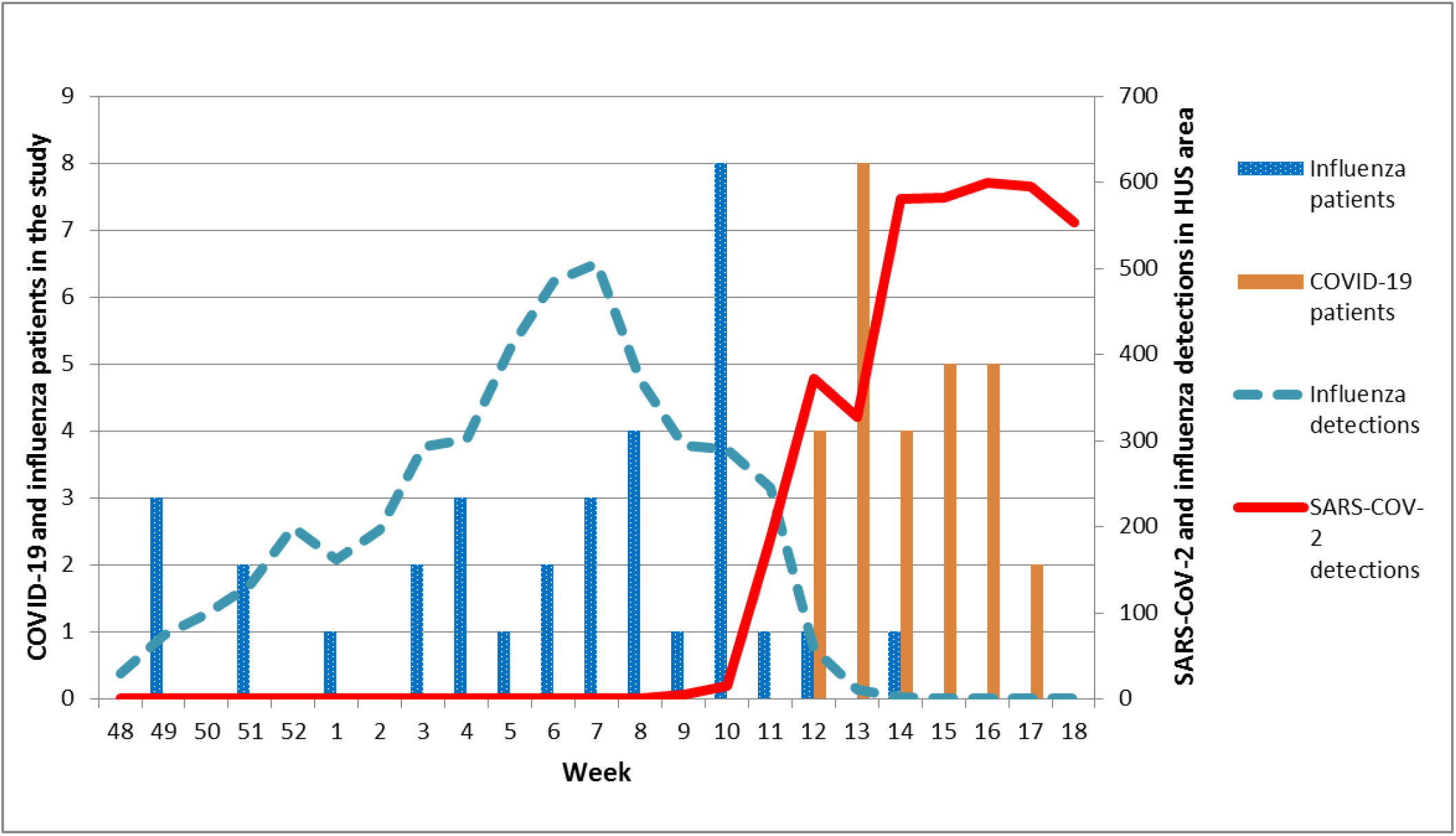
COVID-19 and influenza patients included in the study (bars) and weekly SARS-CoV-2 and influenza findings in the National Infectious Diseases Register (NIDR) in the HUS area (lines) between December 2019 and April 2020. The lockdown put in place to prevent the spread of COVID-19 since week 12 and the decrease in influenza testing likely caused a sudden drop in influenza cases reported to NIDR.

## Materials and methods

### Study design

We conducted a prospective observational study at the HUS Jorvi Hospital, Espoo, Finland together with the Finnish Institute for Health and Welfare (THL) as a part of the international DRIVE collaboration (www.drive-eu.org). The study was primarily focused on estimating influenza vaccine effectiveness and disease burden but its objectives included collecting data on other respiratory pathogens for the needs of national surveillance.

The study was approved by the Local Ethics Committee of the HUS Helsinki University Hospital. Appropriate study permits were obtained from HUS, THL and the municipalities involved. Informed consent was obtained from the patients or the next of kin of those critically ill.

### Study population

HUS Jorvi Hospital, which is a tertiary care hospital of Helsinki University Hospital (HUS), provides specialized care for the 339 000 residents of the cities of Espoo, Kauniainen and Kirkkonummi.

All community-dwelling adult (18+-year-old) SARI patients admitted to the HUS Jorvi Hospital were interviewed by the study nurse and recruited to the study if they fulfilled the eligibility criteria listed in the study protocol and gave their informed consent (Appendix 1). A SARI patient was defined as a hospitalized person with at least one systemic symptom and at least one respiratory symptom. Initially, only patients with symptom onset within the last 7 days were eligible; however, during the COVID-19 epidemic, SARI patients were recruited symptom onset notwithstanding to include all COVID-19 patients.

### Data collection

Data were obtained from patient interviews and electronic medical records by the study nurses and physicians. Comorbidities, smoking, influenza vaccinations, SARI symptoms, triage vitals, routine laboratory test results and radiological images, intensive care unit (ICU) admission, invasive mechanical ventilation and outcomes were collected. Overnight hospitalizations during the past 12 months were collected and verified from the Care Register for Health Care (HILMO). The severity of chronic conditions was assessed using the McCabe score. National Early Warning Score 2 (NEWS2) at admission was calculated. The severity of infection was classified as in WHO interim guidance for COVID-19 [24]. In short, the categories were mild disease, pneumonia, severe pneumonia with respiratory rate > 30 breaths/min, severe respiratory distress, or SpO2 ≤ 93% on room air, and ARDS. In case of readmission to HUS for the same reason within three days of hospital discharge, the hospitalization was reported as one episode.

Routine laboratory test results at admission and highest or lowest values during hospitalization and radiological imaging were extracted. Chest x-ray images were interpreted by a radiologist unaware of the microbiologic diagnosis of the patients. Anemia was defined as hemoglobin < 134 g/l for men and < 117 g/l for women, leukopenia as white blood cells < 3.4 × 10^9^/l and lymphopenia as blood lymphocytes < 1.2 × 10^9^/l and thrombocytopenia as platelet count < 150 × 10^9^/l.

### Laboratory testing for respiratory pathogens

Respiratory samples taken from nasopharynx, oropharynx or trachea for routine diagnostics were tested at the Helsinki University Hospital Laboratory (HUSLAB). Influenza testing was performed using Xpert®Xpress Flu/RSV assay (Cepheid, Sunnyvale, CA, USA). For the detection of SARS-CoV-2 RNA, the specimens were subjected to one the following three methods: 1) the real-time laboratory-developed SARS-CoV-2 RT-PCR based on Corman et al., 2020; 2) Roche Molecular Systems cobas® SARS-Cov-2 assay (Branchburg, NJ, USA); 3) the Amplidiag®COVID-19 assay (Mobidiag, Helsinki, Finland). Influenza virus subtypes or lineages were defined at the THL Expert Unit for Microbiology.

### Statistical analysis

Categorical (described as proportions) and continuous (described as medians and interquartile ranges) variables were compared using Fisher’s exact test and Mann-Whitney’s U-test, respectively. Statistical analyses were performed using SPSS version 26.0 (IBM SPSS Statistics®) and Stata 16.1 (StataCorp, College Station, TX, USA). To analyze predisposing factors predicting the duration of hospitalization, we did a multivariable Cox regression analysis for the hazard of discharge from the hospital. We entered COVID19, age, sex, BMI and comorbidities (including anemia, cancer, cardiovascular disease, diabetes, hypertension, immunosuppression, kidney disease, liver cirrhosis, neurological diseases, pulmonary diseases, rheumatic diseases and stroke) into the Cox analysis and used backward selection using AIC (Akaike information criterion) to choose our final model. The predictors chosen in the final model were COVID19, age, BMI and diabetes.

## Results

Between 2 December 2019 and 30 April 2020, we recruited altogether 213 eligible SARI patients, of whom those 61 patients positive for either SARS-CoV-2 (28) or influenza (33) by RT-PCR were included in this study. Of the influenza patients, 29 (88%) were positive for influenza A(H1N1)pdm09, 2 (6%) were positive for influenza A(H3N2) and 2 (6%) for influenza B/Victoria. The median ages of COVID-19 and influenza patients were 53 (range 18-81) and 56 (19-87), and the majority were men (17 [61%] vs 22 [67%]) (Table 1).

**Table 1.**
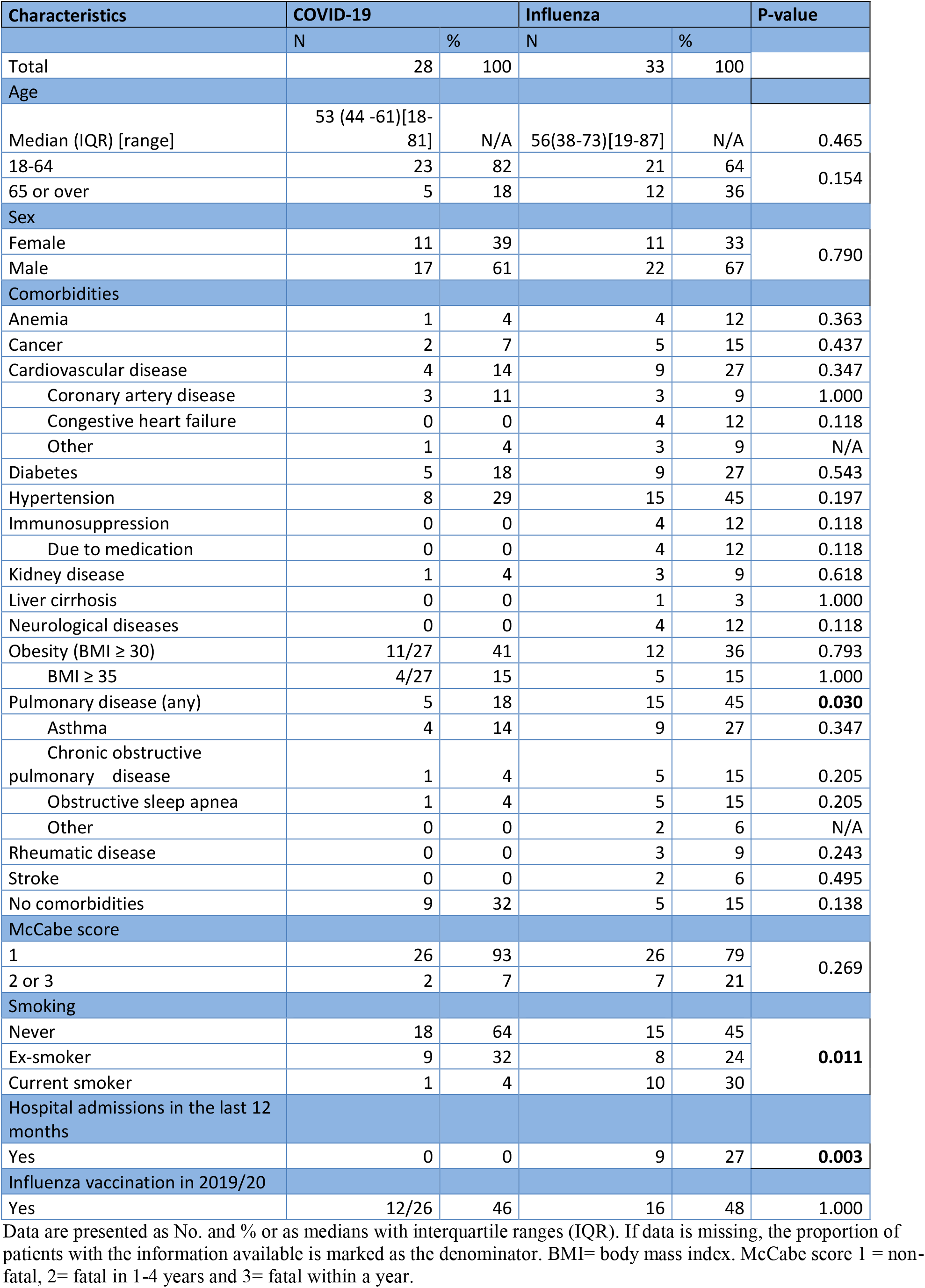
Characteristics of COVID-19 and influenza patients.

| Characteristics | COVID-19 |  | Influenza |  | P-value |
| --- | --- | --- | --- | --- | --- |
|  | N | % | N | % |  |
| Total | 28 | 100 | 33 | 100 |  |
| Age |  |  |  |  |  |
| Median (IQR) [range] | 53 (44 -61)[18-81] | N/A | 56(38-73)[19-87] | N/A | 0.465 |
| 18-64 | 23 | 82 | 21 | 64 | 0.154 |
| 65 or over | 5 | 18 | 12 | 36 |  |
| Sex |  |  |  |  |  |
| Female | 11 | 39 | 11 | 33 | 0.790 |
| Male | 17 | 61 | 22 | 67 |  |
| Comorbidities |  |  |  |  |  |
| Anemia | 1 | 4 | 4 | 12 | 0.363 |
| Cancer | 2 | 7 | 5 | 15 | 0.437 |
| Cardiovascular disease | 4 | 14 | 9 | 27 | 0.347 |
| Coronary artery disease | 3 | 11 | 3 | 9 | 1.000 |
| Congestive heart failure | 0 | 0 | 4 | 12 | 0.118 |
| Other | 1 | 4 | 3 | 9 | N/A |
| Diabetes | 5 | 18 | 9 | 27 | 0.543 |
| Hypertension | 8 | 29 | 15 | 45 | 0.197 |
| Immunosuppression | 0 | 0 | 4 | 12 | 0.118 |
| Due to medication | 0 | 0 | 4 | 12 | 0.118 |
| Kidney disease | 1 | 4 | 3 | 9 | 0.618 |
| Liver cirrhosis | 0 | 0 | 1 | 3 | 1.000 |
| Neurological diseases | 0 | 0 | 4 | 12 | 0.118 |
| Obesity (BMI ≥ 30) | 11/27 | 41 | 12 | 36 | 0.793 |
| BMI ≥ 35 | 4/27 | 15 | 5 | 15 | 1.000 |
| Pulmonary disease (any) | 5 | 18 | 15 | 45 | **0.030** |
| Asthma | 4 | 14 | 9 | 27 | 0.347 |
| Chronic obstructive pulmonary disease | 1 | 4 | 5 | 15 | 0.205 |
| Obstructive sleep apnea | 1 | 4 | 5 | 15 | 0.205 |
| Other | 0 | 0 | 2 | 6 | N/A |
| Rheumatic disease | 0 | 0 | 3 | 9 | 0.243 |
| Stroke | 0 | 0 | 2 | 6 | 0.495 |
| No comorbidities | 9 | 32 | 5 | 15 | 0.138 |
| McCabe score |  |  |  |  |  |
| 1 | 26 | 93 | 26 | 79 | 0.269 |
| 2 or 3 | 2 | 7 | 7 | 21 |  |
| Smoking |  |  |  |  |  |
| Never | 18 | 64 | 15 | 45 | **0.011** |
| Ex-smoker | 9 | 32 | 8 | 24 |  |
| Current smoker | 1 | 4 | 10 | 30 |  |
| Hospital admissions in the last 12 months |  |  |  |  |  |
| Yes | 0 | 0 | 9 | 27 | **0.003** |
| Influenza vaccination in 2019/20 |  |  |  |  |  |
| Yes | 12/26 | 46 | 16 | 48 | 1.000 |

Data are presented as No. and % or as medians with interquartile ranges (IQR). If data is missing, the proportion of patients with the information available is marked as the denominator. BMI= body mass index. McCabe score 1 = non-fatal, 2= fatal in 1-4 years and 3= fatal within a year.

At least one comorbidity was present in 19 (68%) of COVID-19 and 28 (85%) of influenza patients (Table 1). In COVID-19 patients, the most common comorbidities were obesity (11, 41%) and hypertension (8, 29%) whereas in influenza patients hypertension and pulmonary diseases were most prevalent (15, 45% respectively). Pulmonary diseases and current smoking were significantly less common among COVID-19 than influenza patients (5 [18%] vs 15 [45%], P=0.03 and 1 [4%] vs10 [30%], P=0.008). Fatal comorbidities (McCabe score 2 or 3) were slightly less prevalent among COVID-19 than influenza patients (2 [7%] and 7 [21%], P=0.160). None of the COVID-19 patients had been previously hospitalized during the last 12 months compared with 9 (27%) of influenza patients (P = 0.003).

At admission, the clinical picture of COVID-19 was similar to that of influenza with reported fever and cough being the most common symptoms. Headache was more common among COVID-19 patients (22/25, 85% vs 16/31, 52%, P=0.004). COVID-19 patients were hospitalized on average on day 10 of symptoms compared with day 3 for influenza patients (Table 2).

**Table 2.** Characteristics and laboratory and radiological findings of COVID-19 and influenza patients.

| Characteristics and findings | COVID-19 |  | Influenza |  | P-value |
| --- | --- | --- | --- | --- | --- |
|  | No. | % | No. | % |  |
| Total | 28 | 100 | 33 | 100 |  |
| Days from symptom onset to hospitalization |  |  |  |  |  |
| Duration of symptoms at hospitalization, median days (IQR) [range] | 10 (7-12) [1-23] | N/A | 3 (2-5) [1-8] | N/A | **<0.001** |
| 0-3 days | 4 | 14 | 25 | 76 |  |
| 4-7 days | 6 | 21 | 8 | 24 |  |
| ≥ 8 days | 18 | 64 | N/A | N/A |  |
| Reported symptoms |  |  |  |  |  |
| Fever ≥ 38 | 28 | 100 | 33 | 100 | N/A |
| Headache | 22/25 | 88 | 16/31 | 52 | **0.004** |
| Myalgia | 15/25 | 60 | 15/31 | 48 | 0.430 |
| Deterioration of general condition | 27 | 96 | 32 | 97 | 1.000 |
| Cough | 27 | 96 | 30 | 91 | 0.618 |
| Sore throat | 8/24 | 33 | 17/32 | 53 | 0.179 |
| Dyspnea | 26 | 93 | 29 | 88 | 0.678 |
| Sudden onset | 11/23 | 48 | 19/31 | 61 | 0.410 |
| Clinical characteristics at admission, median (IQR) |  |  |  |  |  |
| Temperature | 38.4 (37.5-38.9) | N/A | 38.4 (37.6-39.4) | N/A | 0.482 |
| Fever ≥ 38 | 17 | 61 | 22 | 67 | 0.790 |
| Respiratory rate | 22 (18-28) | N/A | 20 (18-24) | N/A | 0.401 |
| Oxygen saturation on room air | 93 (90-94) | N/A | 94 (90-96) | N/A | 0.616 |
| NEWS2 score (IQR) [range] | 5 (3-6)[0-8] | N/A | 4 (2-6)[0-10] | N/A | 0.423 |
| Laboratory results at admission, median (IQR) |  |  |  |  |  |
| Hemoglobin, g/l (ref. range 134-167 men, 117-155 women) | 142 (129-149) | N/A | 134 (123-145) | N/A | 0.078 |
| Anemia | 4 | 14 | 10 | 30 | 0.222 |
| Leukocyte count, x 10<sup>9</sup>/L (reference range 3.4-8.2) | 5.1 (4.0-6.3) | N/A | 6.7 (5.4-10.9) | N/A | **0.002** |
| Leukopenia | 3/27 | 11 | 1 | 3 | 0.318 |
| Leukocytosis | 3/27 | 11 | 13 | 39 | **0.019** |
| Platelet count, x 10<sup>9</sup>/L (150-360) | 201 (129-269) | N/A | 182 (164-218) | N/A | 0.538 |
| Thrombocytopenia < 150 x 10<sup>9</sup>/L | 11 | 39 | 4 | 12 | **0.019** |
| C-reactive protein, mg/L (<4) (IQR) [range] | 55 (39-119) [12-186] | N/A | 57 (28-117) [7-214] | N/A | 0.965 |
| Creatinine, μmol/l (60-100 men, 50-90 women) (IQR)[range] | 75 (57-87) [42-740] | N/A | 77 (66-88) [40-557] | N/A | 0.406 |
| Alanine aminotransferase, U/L (< 50 men, < 30 women) (IQR)[range] | 42 (30-75) [19-127] | N/A | 23 (18-47) [12-123] | N/A | **0.011** |

| Laboratory findings during hospitalization, median (IQR) |  |  |  |  |  |
| --- | --- | --- | --- | --- | --- |
| Anemia | 7 | 25 | 11 | 33 | 0.578 |
| Leukopenia | 6 | 21 | 4 | 12 | 0.490 |
| Lymphopenia | 18/25 | 72 | 11/19 | 58 | 0.357 |
| Thrombocytopenia < 150 x 10<sup>9</sup>/L | 11 | 39 | 9 | 27 | 0.414 |
| Highest C-reactive protein, mg/L [range] | 159 (90-239) [12-410] | N/A | 79 (42-171) [8-289] | N/A | **0.019** |
| Highest creatinine, µmol/l [range] | 76 (60-89) [45-740] | N/A | 80 (66-91) [47-731] | N/A | 0.482 |
| Highest alanine aminotransferase, U/L [range] | 66 (42-190) [19-457] | N/A | 23 (18-52) [12-140] | N/A | **<0.001** |
| Radiological findings in chest x-ray at admission |  |  |  |  |  |
| No findings | 5 | 18 | 12 | 36 | 0.154 |
| Linear opacities | 4 | 14 | 14 | 42 | **0.024** |
| Bilateral basal | 3 | 11 | 14 | 42 |  |
| Bilateral peripheral | 1 | 4 | 0 | 0 |  |
| Ground glass opacities/Consolidation | 19 | 68 | 7 | 21 | **<0.001** |
| Unilateral or local | 0 | 0 | 2 | 6 |  |
| Bilateral basal | 8 | 29 | 4 | 12 |  |
| Bilateral perihilar | 0 | 0 | 1 | 3 |  |
| Bilateral peripheral | 9 | 32 | 0 | 0 |  |
| Bilateral diffuse | 2 | 7 | 0 | 0 |  |

Data are presented No. and % or as medians with interquartile ranges (IQR). If data is missing, the proportion of patients with the information available is marked as the denominator when applicable. Leukocyte count was missing from one COVID-19 patient at admission. ALAT was taken only from 25/28 of COVID-19 and 19/33 of influenza patients.

The laboratory results of COVID-19 and influenza patients that differed significantly at admission included leukocyte and platelet counts and alanine aminotransferase (ALAT)(Table 2). CRP values were similar at admission but rose significantly higher in COVID-19 patients during hospitalization. ALAT was higher among COVID-19 patients than influenza patients both at admission and during hospitalization. Blood cultures were taken from 23/28 of COVID-19 and 32/33 of influenza patients and were negative.

In chest x-rays at admission, ground-glass opacities (GGO) and consolidations were observed significantly more often in the images of COVID-19 than influenza patients (19 [68%] and 7 [21%], P < 0.001), whereas linear opacities were more common among influenza patients (Table 2). Typically the GGO and consolidations present in the images of COVID-19 patients were bilateral and peripheral or basal (Figure 2). No findings were observed in 5 (18%) of COVID-19 and 12 (36%) of influenza patients (P=0.154). Three of the five COVID-19 patients with no radiological findings at admission developed consolidations in chest x-ray within one week.

**Figure 2:**
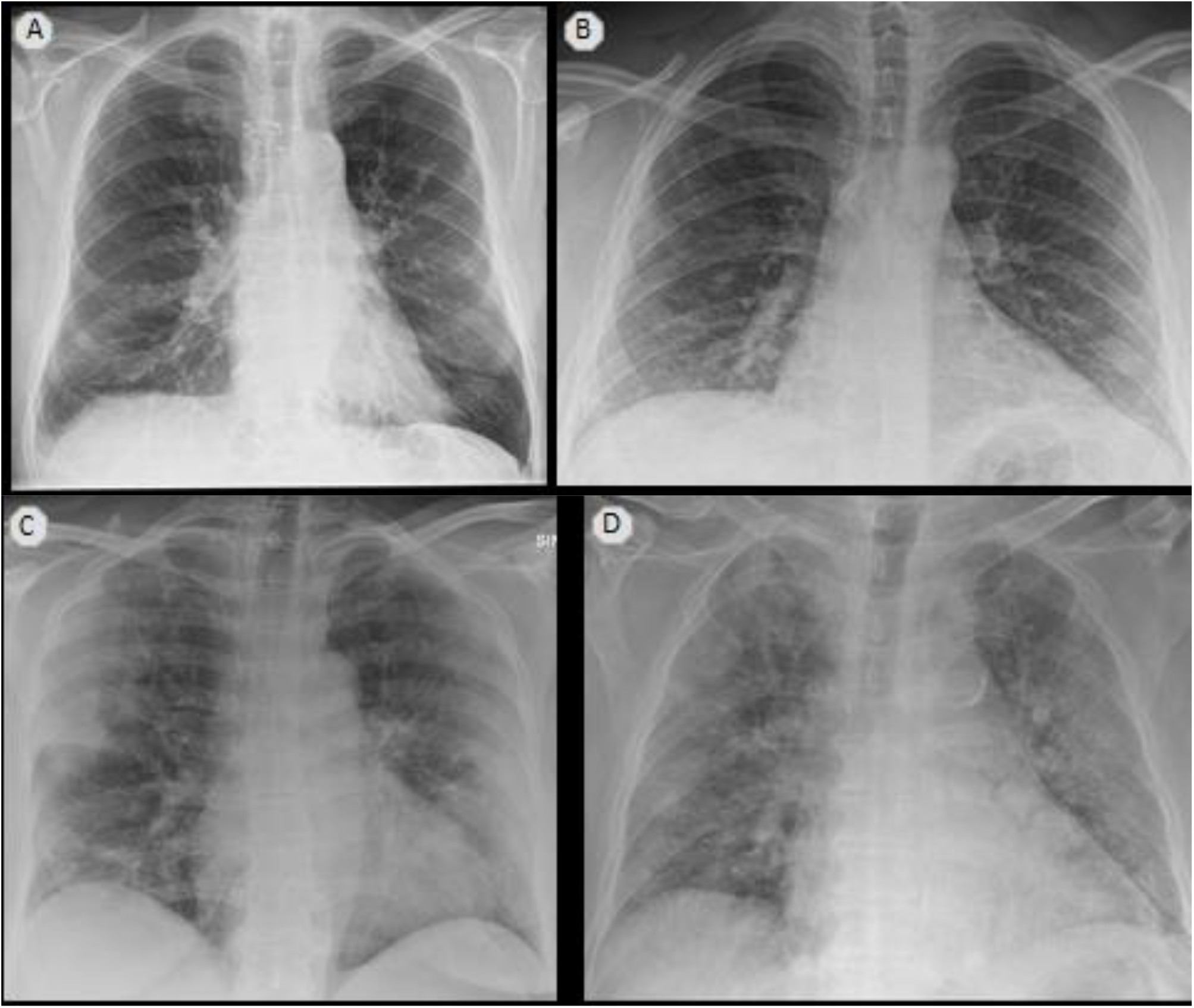
Chest x-rays of COVID-19 and influenza patients at admission. (A) An 88-year-old man, influenza, day 6 of symptoms: bilateral basal linear opacities (B) A 28-year-old man with COVID-19, day 9 of symptoms, bilateral basal ground-glass opacities (GGO) (C) A 51-year-old woman with COVID-19, day 13 of symptoms: bilateral peripheral GGO and consolidations. (D) A 68-year-old man with COVID-19, the third day of symptoms: bilateral diffuse GGO and consolidations.

Severe disease (severe pneumonia or ARDS) and ICU admission were significantly more common among COVID-19 patients than influenza patients (26 [93%] vs 19 [58%], P=0.003 and 8 [29%] vs 2 [6%], P=0.034)(Table 3). Notably, all patients admitted to the ICU were men. The median age of the eight COVID-19 patients admitted to the ICU was 60.5 (range 52-61) while the two influenza patients were 38 and 62 years old. All eight COVID-19 patients developed ARDS and were mechanically ventilated compared with one of the two influenza patients and their median stay at the ICU was 20 (range 11-41) compared with 9.5 (3-16) days. Two COVID-19 patients were at the ICU twice and both episodes were included in the total duration of their ICU stay.

**Table 3.**
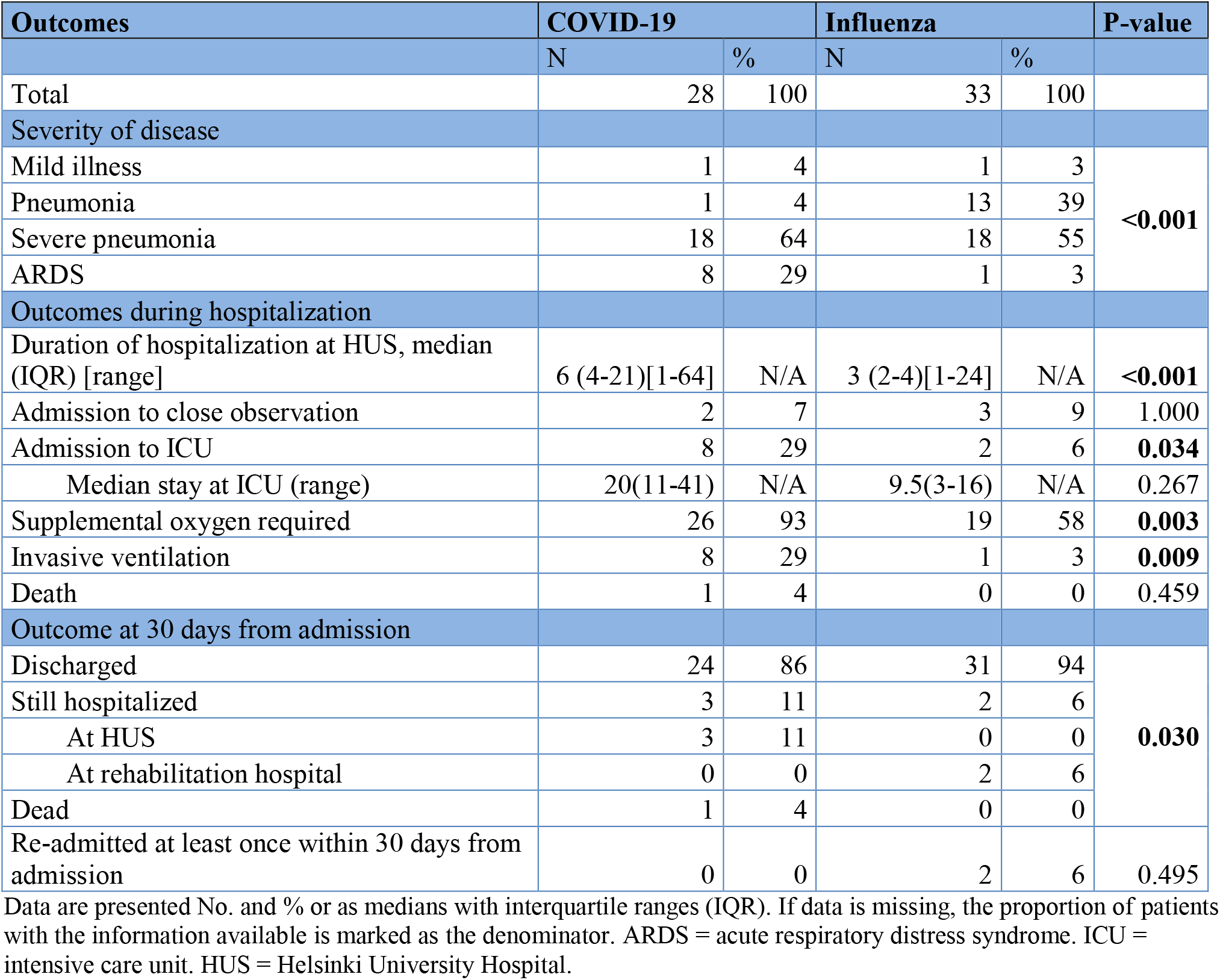
Outcomes of COVID-19 and influenza patients.

As of 5 June 2020, 60 of the 61 patients had been discharged from HUS Jorvi Hospital and one COVID-19 patient had died. COVID-19 patients were hospitalized significantly longer than influenza patients (6 days [IQR 4-21] vs 3 days [2-4], P<0.001) (Table 3, Figure 3). To further examine whether there were other factors predicting the duration of hospitalization besides COVID-19, we did a multivariable Cox regression analysis. The predictors associated with a longer duration of hospitalization were COVID-19 (hazard ratio [HR] 0.221, 95% CI 0.118, 0.416, P < 0.001), age (HR 0.972, CI 0.955, 0.990, P=0.002), BMI (HR 0.950, CI 0.916, 0.986, P=0.006) and diabetes (HR 0.539, CI 0.272, 1.066, P=0.076).

**Figure 3.**
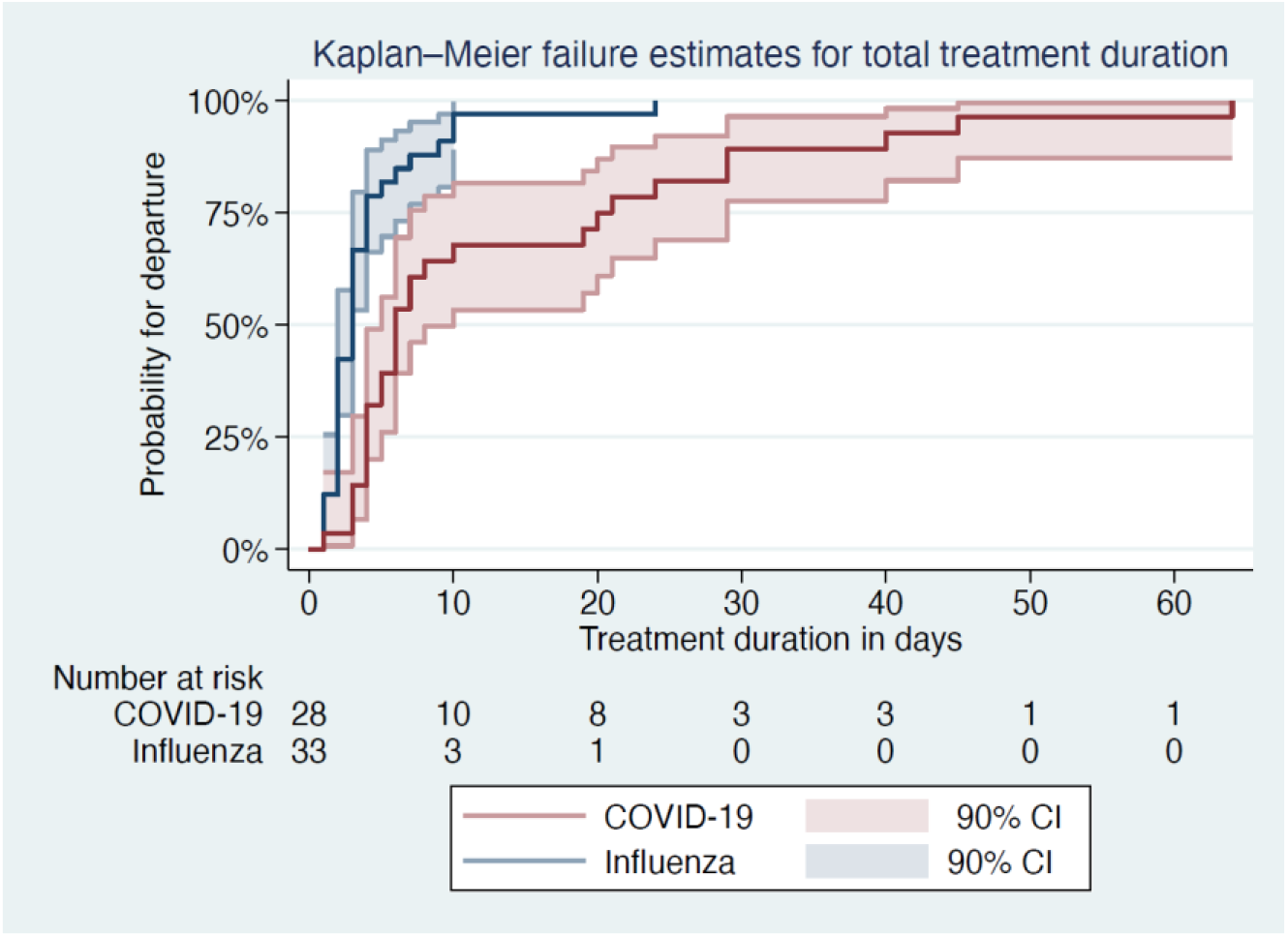
Kaplan-Meier curves for the duration of hospitalization among COVID-19 and influenza patients.

## Discussion

To our knowledge, our study is the first prospective study comparing the clinical characteristics and outcomes of hospitalized adult patients with COVID-19 or seasonal influenza with clinical pictures ranging from mild disease to ARDS. We observed that pulmonary diseases, smoking and previous hospital admissions were less common among COVID-19 than influenza patients. At admission, their symptoms, disease severity and laboratory findings were similar but chest x-ray findings differed. COVID-19 patients developed a more serious disease and required longer hospitalization.

COVID-19 patients had no previous hospitalizations during the past 12 months, 32% had no comorbidities compared with 15% of influenza patients and their comorbidities were less serious based on their McCabe score; however the latter two findings did not reach statistical significance. Together these findings imply that COVID-19 patients were previously healthier than influenza patients.

The lower prevalence of pulmonary diseases among COVID-19 patients was not limited to smoking-related COPD but was also seen for asthma and obstructive sleep apnea. Whether this relates to influenza viruses affecting bronchi more than SARS-CoV-2 for example due to different viral receptors (ACE2 for SARS-CoV-2 and sialic acid-containing molecules for influenza) and viral tropism, remains a subject for further investigation [1,12,25].

Influenza patients were hospitalized earlier after symptom onset than those with COVID-19 as in previous publications on hospitalized A(H1N1)pdm09 patients [26]. Fever, cough and dyspnea were the most common symptoms in both groups. Differences in laboratory findings at admission were not remarkable enough to enable differential diagnostics based on laboratory findings alone. As in previous studies, laboratory findings associated with COVID-19 included lymphocytopenia, thrombocytopenia and elevated liver enzymes [9,10,17,23]. During hospitalization, CRP values rose significantly higher among COVID-19 patients, especially in patients admitted to ICU.

Of the COVID-19 patients, 18% had no radiological abnormalities in chest x-ray at admission compared with 41% in a previous study, which is likely explained by the bigger proportion of severe disease in our study [10]. In line with previous CT findings, bilateral basal or peripheral ground-glass opacities (GGO) and consolidations were typically seen in the chest x-rays of COVID-19 patients, which may help in differential diagnostics. In chest CTs of COVID-19 patients, initially unilateral and multifocal GGO evolving to diffuse bilateral GGO and later consolidations in the course of the infection have been described [27]. Rounded opacities, interlobular septal thickening and peripheral distribution and GGO in the CTs of ARDS patients have been more common among COVID-19 than influenza patients [28,29].

Despite similar clinical presentation at admission, COVID-19 patients developed a more severe disease during hospitalization and required longer hospitalization than influenza patients. In multivariable Cox regression analysis, also increasing age, higher BMI and diabetes were associated with longer hospitalization. In previous studies, along with age and other comorbidities, obesity has been a risk factor for severe disease and ICU admission in both COVID-19 and influenza, which supports our findings [30,31].

Of the COVID-19 patients included in our study, 29% were admitted to the ICU and 4% died compared with 6% and 0% of influenza patients. In a recent large study from the USA, 14.2% of hospitalized COVID-19 patients were admitted to ICU and 21% died [32]. These differences may partially be explained by the exclusion of institutionalized and elderly patients with poor prognosis from our study. Our findings are aligned with the figures from whole HUS, where 629 COVID-19 patients had been hospitalized by 11 June 2020, 139 (22%) of them had been admitted to ICU and 38 (6%) had died (Personal communication from HUS).

In a previous study, COVID-19 patients with ARDS were older, had lower severity of illness scores at presentation and lower Sequential Organ Failure Assessment (SOFA) score-adjusted mortality than H1N1 patients with ARDS (29). In our study there were too few ARDS patients for similar comparisons but ARDS developed more often among COVID-19 patients (29% vs 3%).

Our study setting offered several advantages. We included COVID-19 patients since the beginning of the outbreak in Finland. All SARI-patients were systematically interviewed and recruited usually before their COVID-19 or influenza test results were available, minimizing selection bias. Data were verified from several sources. Patients had known outcomes as the follow-up lasted until the last patient was discharged. The community-dwelling working-age population of Espoo, the second largest city of Finland, is well represented as HUS Jorvi Hospital provides their specialized care. All patients were recruited during the same influenza season and 88% had influenza A(H1N1)pdm09, the offspring of the pandemic A(H1N1) of 2009 currently circulating as seasonal influenza. Thus, the variation in the clinical course of influenza due to different circulating influenza strains or different composition of seasonal influenza vaccination was minimized.

Our study had several limitations. The sample size was small, which should be taken into account when interpreting the results. All patients admitted to HUS Jorvi Hospital with COVID-19 or influenza were not eligible; for example they lived outside of the Jorvi Hospital catchment area or did not consent to participate. Elderly patients were underrepresented as patients with pre-existing treatment limitations or not requiring specialized care were admitted to the secondary care Espoo Hospital. The time from symptom onset to hospitalization may not be completely comparable between COVID-19 and influenza patients because, until March, only patients with symptom onset within the last 7 days were included. Furthermore, this study is based on clinical data and no extra laboratory testing or radiological imaging was done in addition to the detection of influenza and SARS-CoV-2 RNA. Consequently, CT scans, coagulation markers, cardiac biomarkers or blood gas analyses were mostly not available and thus were not included here. During the COVID-19 epidemic, occasionally SARS-CoV-2 detection alone was conducted without influenza testing on clinical grounds, however, of the COVID-19 patients included in this study, 21 (75%) were tested for influenza and no co-infections were found.

In conclusion, COVID-19 requiring hospitalization had a more severe clinical course than influenza. As SARS-CoV-2 and influenza may continue to circulate simultaneously and influenza already strains health care capacity every influenza season, this has important implications for public health policies for COVID-19 mitigation. Larger studies are needed to further examine the differences between COVID-19 and influenza.

## Data Availability

The data are available from the corresponding author upon reasonable request respecting GDPR and with permission from HUS and THL.

## Acknowledgements

Thank you to our study nurses Outi Debnam and Eija Mikkola for their important work in recruiting the patients to this study and to all patients who participated. We are also very thankful to the DRIVE collaboration for providing the funding and support on which to build our study. We gratefully thank Riitta Santanen, Päivi Hirttiö-Tallbacka, Alena Kaijalainen, Minna Haanpää and Johanna Mustajoki for their expert technical assistance and Esa Ruokokoski for data management.

## Declarations

### Ethics approval

This study was performed in line with the principles of the Declaration of Helsinki. Approval was granted by the Local Ethics Committee of the Helsinki University Hospital (Date 3 October 2019, case number: HUS/2517/2018)

### Consent to participate

Informed consent was obtained from all study participants or, in the case of the critically ill patients unable to give their consent personally, from their next of kin.

### Consent for publication

Informed consent included consent to publish study data. Additional consent for the publication of chest x-rays was obtained.

### Availability of data and material

The data that support the findings of this study are available from the corresponding author upon reasonable request respecting GDPR and with permission from HUS and THL.

### Code availability

Not applicable.

### Author contributions

All authors have made substantial contributions to this work and have approved the final manuscript.

Concept and design: RS, RA, HN, KS, RL, NI

Acquisition, analysis, or interpretation of data: RA, JM, TK, KS, JO, NI, AH, RL, SK

Resources: HN, RS, RA, KS, NI, RL

Writing original draft: RA

Writing: review and editing: RA, HN, KS, SK, JO, TK, RS

Supervision: HN, RS and KS

